# ASSESSING THE RELATIONSHIP BETWEEN MONOALLELIC *PARK2* MUTATIONS AND PARKINSON’S RISK

**DOI:** 10.1101/2020.06.26.20138172

**Authors:** Steven J Lubbe, Bernabe Bustos, Jing Hu, Dimitri Krainc, Theresita Joseph, Jason Hehir, Manuela Tan, Weijia Zhang, Valentina Escott-Price, Nigel M Williams, Cornelis Blauwendraat, Andrew B Singleton, Huw R Morris, for International Parkinson’s Disease Genomics Consortium (IPDGC)

## Abstract

Biallelic *PARK2* (Parkin) mutations cause autosomal recessive Parkinson’s (PD); however, the role of monoallelic *PARK2* mutations as a risk factor for PD remains unclear. We investigated the role of single heterozygous *PARK2* mutations in three large independent case-control cohorts totalling 10,858 PD cases and 8,328 controls. Overall, after exclusion of biallelic carriers, single *PARK2* mutations were more common in PD than controls conferring a >1.5-fold increase in risk of PD (P=0.035), with meta-analysis (19,574 PD cases and 468,488 controls) confirming increased risk (OR=1.65, P=3.69E-07). Carriers were shown to have significantly younger ages at onset compared to non-carriers (NeuroX: 56.4 *vs*. 61.4 years; Exome: 38.5 *vs*. 43.1 years). Stratifying by mutation type, we provide preliminary evidence for a more pathogenic risk profile for single *PARK2* copy number variant (CNV) carriers compared to single nucleotide variant carriers. Studies that did not assess biallelic *PARK2* mutations or consist of predominantly early-onset cases may be biasing these estimates, and removal of these resulted in a loss of association (OR=1.23, P=0.614; n=4). Importantly, when we looked for additional CNVs in 30% of PD cases with apparent monoallellic *PARK2* mutations we found that 44% had biallelic mutations suggesting that previous estimates may be influenced by cryptic biallelic mutation status. While this study supports the association of single *PARK2* mutations with PD, it highlights confounding effects therefore caution is needed when interpreting current risk estimates. Together, we demonstrate that comprehensive assessment of biallelic mutation status is essential when elucidating PD risk associated with monoallelic *PARK2* mutations.

## INTRODUCTION

Parkinson’s (PD) is a multifactorial neurodegenerative disease. Common variation within 78 independent loci increase PD risk (1). Pathogenic mutations in autosomal dominant genes (*LRRK2, SNCA* and *VPS35*) as well as biallelic mutations in autosomal recessive (AR) genes (*PARK2, DJ-1, PINK1* and *FBXO7*) cause Mendelian PD (2). It has been suggested that single heterozygous pathogenic AR mutations can increase the risk of PD, and several lines of evidence have been provided for and against mutations (reviewed in Klein *et al*., 2007) (3). Previous studies may have been confounded by differences in methods for mutation detection in cases and controls. Biallelic AR mutations in PD genes are rare in PD cases, but single heterozygous mutations in specific AR PD genes are more common and are estimated, depending on the population, to occur in between 0.6% and 3% of unaffected control individuals (4–7). Accurate estimation of any risk associated with single heterozygous AR mutations is therefore essential for the counselling of biallelic carriers, monoallelic carriers and their family members. Furthermore, understanding the risk associated with single AR mutations may provide important insights into disease biology. Here, we investigate whether single carriers of disease-causing *PARK2* mutations are at an increased risk for PD using three large independent case-control cohorts using exome-focused genotype data, whole exome sequencing and resequencing data from the International Parkinson’s Disease Genomics Consortium (IPDGC).

## RESULTS

We identified a total of 109 monoallelic *PARK2* mutation carriers in 12,251 PD cases and controls (72 PD, 37 controls), carrying 19 different *PARK2* variants known to cause AR PD in the biallelic state, using the NeuroX genotyping platform (8). It is possible that the identified PD cases represent misclassified true biallelic *PARK2* PD cases. To confirm whether PD cases carry a single pathogenic allele or whether a second variant was missed, we (i) reviewed diagnostic reports if available (n=4), or (ii) assessed available samples using multiplex ligation-dependent probe amplification (MLPA, n=29). Of the 33 available NeuroX samples, representing ∼30% of our putative monoallelic individuals (5 controls, 13.5%; 28 PD-Monoallelic, 38.9%), six cases (18% of the available samples, 21% of available PD cases) were found to harbour a second mutation and therefore were removed, leaving a total of 66 PD cases for all subsequent analyses (no controls were found to harbour a second mutation).

After removal of cases with established Mendelian biallelic mutations across all known PD genes, 1.0% (66/6,552) of PD cases were found to harbour single heterozygous *PARK2* PD-causing mutations (either heterozygous copy number variants [CNVs] or single nucleotide variants [SNVs], Table 1), compared to 0.6% (37/5,693) of controls. Single heterozygote mutations might increase PD risk (OR=1.55; 95% CI:1.03, 2.33; P=0.035), although ∼70% of putative monogenic cases were not assessed for a second mutation. If the remaining apparent monoallelic cases had a similar rate of occult biallelic mutations, then the true underlying monoallelic carrier rate could be estimated to be lower at 0.9%, and there would be no difference between cases and controls (OR=1.39; 95% CI:0.90, 2.16; P=0.117). Using age at onset (AAO) data from 5,710 (87.1%) cases, we found that NeuroX PD cases with single *PARK2* mutations have significantly lower AAOs (Average=56.4 years) than cases without known mutations (Average=61.4 years; Coeff=-5.04; 95% CI:-8.32, −1.71; P=0.003).

**Table 1:**
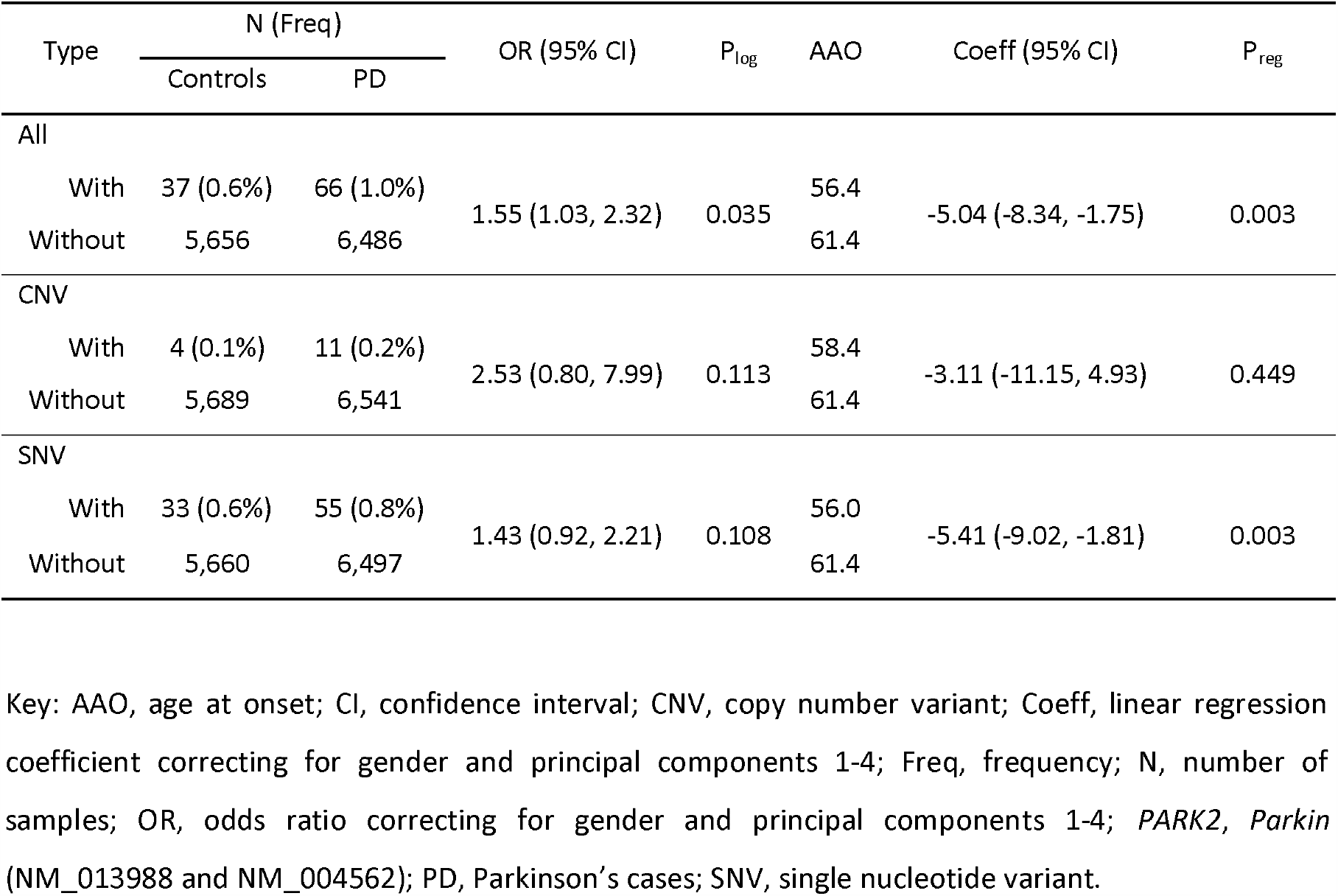
NeuroX Parkinson’s risk profiles associated with single heterozygous *PARK2* mutations.

| Type | N (Freq) |  | OR (95% CI) | P<sub>log</sub> | AAO | Coeff (95% CI) | P<sub>reg</sub> |
| --- | --- | --- | --- | --- | --- | --- | --- |
|  | Controls | PD |  |  |  |  |  |
| All |  |  |  |  |  |  |  |
| With | 37 (0.6%) | 66 (1.0%) | 1.55 (1.03, 2.32) | 0.035 | 56.4 | -5.04 (-8.34, -1.75) | 0.003 |
| Without | 5,656 | 6,486 |  |  | 61.4 |  |  |
| CNV |  |  |  |  |  |  |  |
| With | 4 (0.1%) | 11 (0.2%) | 2.53 (0.80, 7.99) | 0.113 | 58.4 | -3.11 (-11.15, 4.93) | 0.449 |
| Without | 5,689 | 6,541 |  |  | 61.4 |  |  |
| SNV |  |  |  |  |  |  |  |
| With | 33 (0.6%) | 55 (0.8%) | 1.43 (0.92, 2.21) | 0.108 | 56.0 | -5.41 (-9.02, -1.81) | 0.003 |
| Without | 5,660 | 6,497 |  |  | 61.4 |  |  |

Key: AAO, age at onset; CI, confidence interval; CNV, copy number variant; Coeff, linear regression coefficient correcting for gender and principal components 1-4; Freq, frequency; N, number of samples; OR, odds ratio correcting for gender and principal components 1-4; *PARK2*, *Parkin* (NM\_013988 and NM\_004562); PD, Parkinson's cases; SNV, single nucleotide variant.

We next sought to explore the potential increased risk in two independent IPDGC case-control cohorts, using exome sequencing (cases=1,235; controls=473) (8) and resequencing data (cases=3,071; controls=2,162). We identified 28 (23 cases, 5 controls) and 52 (36 cases, 16 controls) carriers of single *PARK2* mutations in the exome (as previously described (8)) and resequencing data respectively. CNVs were not determined in the primary exome or resequencing dataset. In the exome cohort, 1.9% (23/1,231) of cases and 1.1% (5/473) of controls, and in the resequencing cohort, 1.2% (36/3,071) of cases and 0.7% (16/2,162) of controls harboured single *PARK2* SNVs. Before searching for occult second mutations, a meta-analysis of the three IPDGC (NeuroX, exome sequencing and resequencing) cohorts revealed a significant ∼1.5-fold increased risk (OR=1.57; 95% CI:1.15, 2.16; P=0.005; I^2^=0.0%, P _het_ =0.960) associated with *PARK2* mutations (Figure 1). AAO data was available on 1,130 PD exome cases (91.8%) and 2,599 resequencing cases (84.6%). Albeit non-significant, exome PD cases carrying single *PARK2* SNVs had lower AAO compared non-carriers (Average=38.5 years *vs*. 43.1 years; Coeff=-4.34; 95% CI:-8.95, 0.28; P=0.066), with carriers having significantly lower AAO in resequencing cases (Average=52.6 years *vs*. 60.5 years; Coeff=--7.84; 95% CI:-12.59, −3.09; P=0.001). We then used MLPA to search for potentially missed *PARK2* CNVs in mutation carriers. Four of the nine available exome DNA samples (44%, all PD cases) were found to harbour a missed second mutation and were removed from subsequent analyses. Assuming a similar rate of occult biallelic carriage across both datasets, the true rate of monoallelic cases could be estimated to be 1.2% and 0.7% in the exome and resequencing cases as compared with 1.1% and 0.7% of controls, respectively.

**Figure 1:**
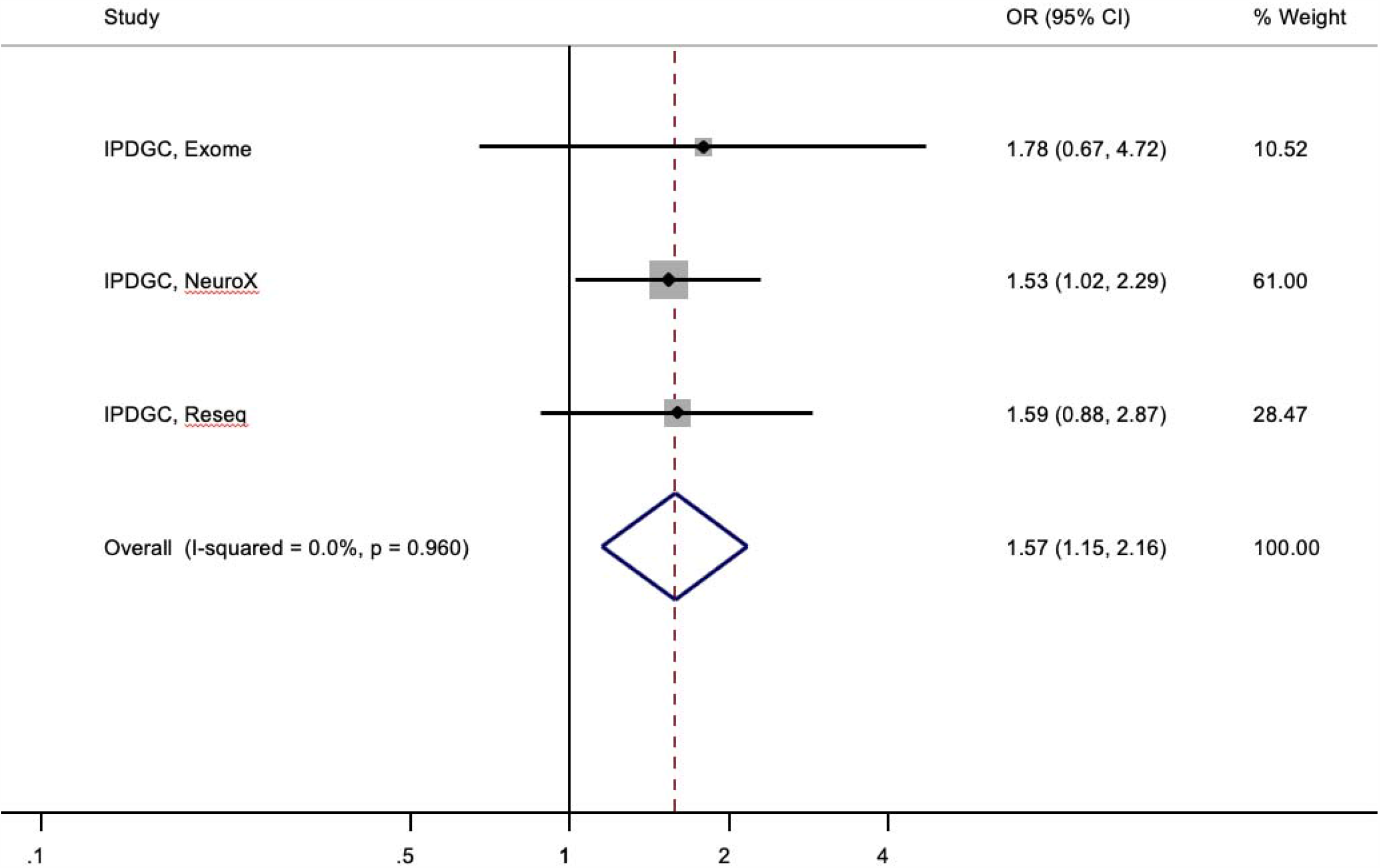
Forest plot of the odds ratio (OR) of the Parkinson’s risk associated with heterozygous *PARK2* mutations in three independent International Parkinson’s disease Genomics Consortium (IPDGC) cohorts. Boxes denote OR point estimates, their areas proportional to the inverse variance weight of the estimate. Horizontal lines represent 95% CIs. Vertical dashed line represents pooled OR point estimates. Key: CI, confidence intervals; PARK2, Parkin (NM_013988 and NM_004562); %, percentage; Reseq, resequencing.

We next performed a meta-analysis of available cohorts and studies that reported heterozygous PD-causing mutation rates in cases and controls, from European ancestry cohorts only. Three cohorts (Parkinson’s Progression Markers Initiative, PPMI, https://www.ppmi-info.org/; UK Biobank Genotyping and Exome cohorts, https://www.ukbiobank.ac.uk/) and 21 published studies were included in our analyses (5,6,9–27). Including our cohorts, the meta-analysis revealed a significant 1.65-fold increased PD risk in single *PARK2* mutation carriers (95% CI: 1.36, 2.00; P=3.69E-07; I^2^=0.0%, P _het_ =0.594) (Figure 2).

**Figure 2:**
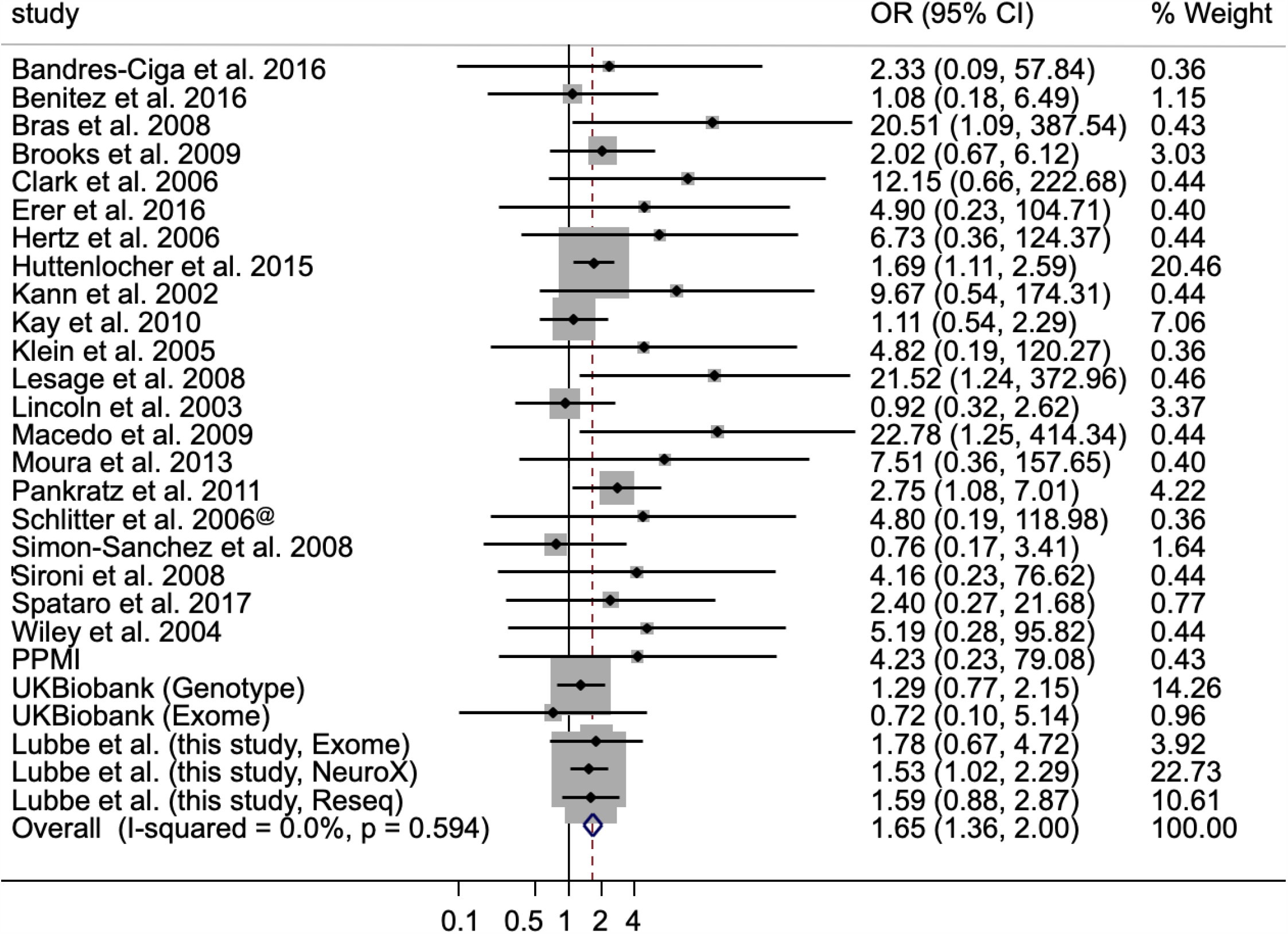
Forest plot of the odds ratio (OR) of the Parkinson’s risk associated with heterozygous *PARK2* mutations. Boxes denote OR point estimates, their areas proportional to the inverse variance weight of the estimate. Horizontal lines represent 95% CIs. Vertical dashed line represents pooled OR point estimates. Key: CI, confidence intervals; PARK2, Parkin (NM_013988 and NM_004562); %, percentage; German samples only.

As in our study, occult second mutations are likely biasing these estimates, so we therefore restricted our meta-analysis to 9 studies (9/21, 43%) that searched for a second *PARK2* mutation (Supplementary Table 1). Based on these studies, single *PARK2* mutations confer a 2-fold increase in PD risk in carriers (OR=2.00, 95% CI:1.10, 3.62, P=0.023; I^2^ =1.3%, P _het_ =0.423) (Supplementary Figure 1).

Inclusion of studies that used predominantly early-onset PD (EOPD) cases may additionally be inflating these estimates, we therefore repeated the meta-analysis excluding these EOPD studies (15/21, 71%), which demonstrated a 1.5-fold significant increased risk in carriers (OR=1.50, 95% CI:1.22, 1.84; P=1.05E-04; I^2^=0%, P _het_ =0.927) (Supplementary Figure 2).

Restricting our analysis to the four non-EOPD studies that searched for biallelic carriers demonstrated that single *PARK2* mutations were not associated with an increased PD risk in these cohorts (OR=1.23, 95% CI:0.55, 2.75, P=0.614; I^2^=0.0%, P _het_ =0.657; Supplementary Figure 3).

**Figure 3:**
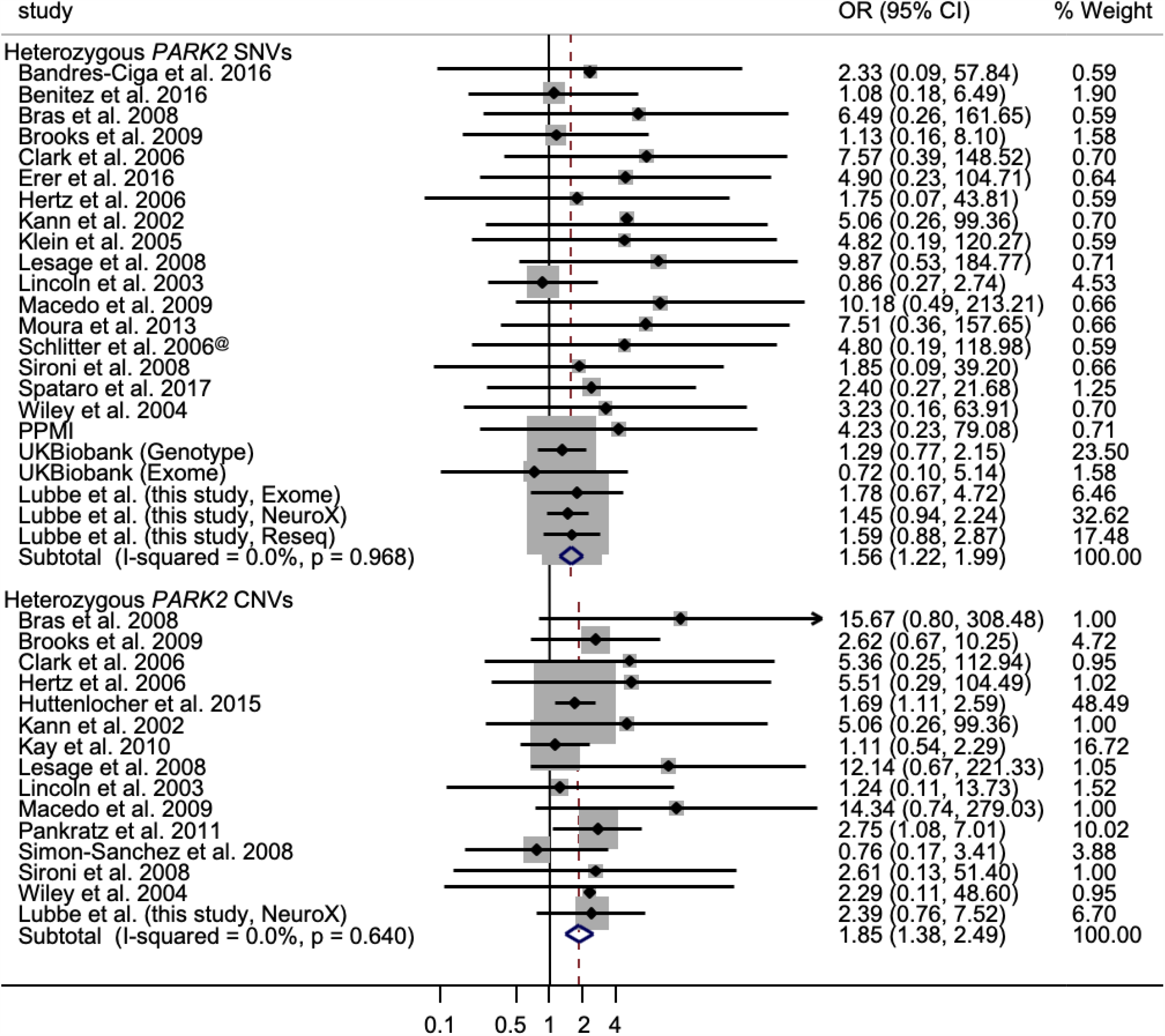
Forest plot of the odds ratio (OR) of the Parkinson’s risk associated with heterozygous *PARK2* single nucleot ide variant (SNV) and copy umber variant (CNV) carriers. Boxes denote OR point estimates, their areas proportional to the inverse variance weight of the estimate. Horizontal lines re present 95% CIs. Vertical dashed line represents pooled OR point estimates. Key: CI, confidence intervals; IPDGC, International Parkins on’s disease Genomics Consortium; PARK2, Parkin (NM_013988 and NM_004562); %, percentage; PPMI, Parkinson’s Progression Markers Initiative; Reseq, Resequencing; German samples only.

The pathogenicity of the common *PARK2* p.R275W variant in AR PD is not as clear cut as other *PARK2* mutations. To test whether the observed SNV association is driven by p.R275W, we repeated the meta-analysis after excluding this variant. Removal of p.R275W resulted in a marginally increased estimate (OR=1.76, 95% CI:1.37, 2.28, P=1.42E-05; I^2^=0.0%, P _het_ =0.673; Supplementary Table 2; Supplementary Figure 4). Limiting our analysis to the 8 cohorts which assessed biallelic mutations indicated a >2-fold increased risk (OR=2.41, 95% CI:1.17, 4.96, P=0.017; I^2^=0.0%, P _het_ =0.791) (Supplementary Figure 5).

The contribution of biallelic *PARK2* CNVs to AR PD is well established; however, that of heterozygous CNV carriers remains unclear. We identified monoallelic *PARK2* CNVs in 0.17% (11/6,552) of non-Mendelian PD cases compared to 0.07% (4/5,693) controls (Table 1) using the NeuroX data only. None of these CNV carriers overlapped with NeuroX SNV carriers. There was a >2.5-fold increase in PD risk for *PARK2* CNV heterozygote carriers compared to controls (OR=2.53; 95% CI:0.80, 7.99; P=0.113) but this was not statistically significant.

It has been suggested that monoallelic *PARK2* CNVs might confer a higher risk that is associated with a more pathogenic profile compared to other AR mutations (28). To assess this, we compared differences in risk between CNV and SNVs carriers in the NeuroX cohort. A total of 55 *PARK2* SNV carriers were seen in non-Mendelian PD cases (55/6,552; 0.8%) compared to 33 controls (33/5,693; 0.6%) (OR=1.43; 95% CI:0.92, 2.21; P=0.108) (Table 1). To test whether *PARK2* CNVs confer a more “pathogenic” risk profile compared to SNVs we performed AAO analysis in the NeuroX data only. Carriers of heterozygous *PARK2* CNVs had a mean AAO of 58.4 years, compared to non-carriers (61.4 years) (Coeff=-3.11; 95% CI:-11.15, 4.93; P=0.449).

To further investigate the potential different risk profiles, we performed separate meta-analyses of published *PARK2* mutation data for SNVs and CNVs. Meta-analyses, including the current data, revealed significant independent increased PD risks for SNVs (OR=1.56, 95% CI:1.22, 2.00, P=4.46E-04; I^2^=0%, P _het_ =0.968) and CNVs (OR=1.85, 95% CI:1.38, 2.50, P=4.55E-05; I^2^=0.0%, P _het_ =0.640) (Figure 3; Supplementary Table 3). Restricting our meta-analysis to studies that searched for second hits suggested that PD risk was larger in carriers of single *PARK2* CNVs (OR=3.11, 95% CI:1.23, 7.89, P=0.016; I^2^=0.0%, P _het_ =0.879) compared to those harbouring heterozygous SNVs (OR=1.59, 95% CI:0.79, 3.20, P=0.191; I^2^=0.0%, P _het_ =0.785).

## DISCUSSION

The role of rare biallelic mutations in *PARK2* in AR PD (MIM#600116) is well established. Here, using data from a large PD case-control cohort, we identified a total of 109 carriers of single heterozygous *PARK2* mutations. After exclusion of PD cases with known mutations, we demonstrated that carriers of single mutations were at a small but significantly increased risk of PD (OR=1.55; 95% CI:1.03, 2.33; P=0.035). This was confirmed by a meta-analysis with two additional IPDGC cohorts (cases=10,954; controls=8,328) which demonstrated a significant >1.5-fold increased risk (P=0.005). Carriers also had significantly lower AAOs than non-carriers (56.4 years *vs*. 61.4 years; P=0.003). Similar findings were seen in the exome and resequencing data for increased risk (Exome, OR=2.20; Resequencing, OR=1.59) and younger AAOs compared to non-carriers (Exome, 38.5 years *vs*. 43.1 years; Resequencing, 52.6 years *vs*. 60.5 years). A meta-analysis of 19,574 PD cases and 468,488 controls from 27 cohorts further confirmed that heterozygous *PARK2* mutations confer an increased PD risk (OR=1.65; P=3.69E-07). However, several confounding factors are likely biasing these estimates in favour of increased risk and are explored below. Large-scale studies in systematically recruited cohorts that have comprehensively interrogated biallelic *PARK2* mutations are therefore needed to accurately determine the risk associated with single mutations.

The relatively common p.R275W (c.823C>T, rs34424986) variant, the most frequent PD-associated variant in *PARK2*, has not been reported in the homozygous state and has only been reported in compound heterozygotes with another mutation in multiple AR PD families (MIM#602544), and has been classified as likely pathogenic. p.R275W reduces protein stability by disrupting binding to phosphorylated ubiquitin and results in reduced Parkin levels (29) supporting the pathogenicity of p.R275W. We examined whether the increased PD risk associated with single *PARK2* variants was driven by this variant. The observation that the OR increases after removal suggests that p.R275W may have reduced effect on enzyme activity compared to other mutations, and that, due to its more common frequency, its presence may be diluting the true effect of heterozygous *PARK2* mutations in PD biology.

Our analysis provides some support for a more “pathogenic” risk profile associated with *PARK2* CNVs as compared with SNVs (CNVs, OR=2.53; SNVs, OR=1.43) in the NeuroX data. Both mutation types appear to be associated with lower AAOs; however, the small number of observed CNVs prevents any definitive conclusions from being drawn. While the meta-analysis results here support the increased risk for *PARK2* CNV carriers (OR=1.85), albeit marginally higher than *PARK2* SNVs (OR=1.56), the increased risk associated with *PARK2* CNVs should be interpreted with care. Small sample sizes, low CNV frequency and failure to investigate/report CNVs may have resulted in an underestimated effect size seen in the meta-analysis. Failure to conclusively look for second *PARK2* hits may also be a potential confounder when trying to estimate the risk associated with single *PARK2* mutations. Additional work in larger cohorts where both *PARK2* SNVs and CNVs are routinely assessed is therefore needed to gain more accurate insight into the different risk profiles associated with different mutation types.

This large study builds on previous work looking at single *PARK2* mutations in PD aetiology. While several failed to identify single known pathogenic *PARK2* mutations in controls thereby supporting increased disease risk, others have found equal frequencies in both cases and controls providing evidence against increased risk (4,7,9,10,24,30–32). These estimates have, however, been based on relatively small sample sets which have made it difficult to conclusively determine if single mutations confer any risk. The inclusion of non-ClinVar (https://www.ncbi.nlm.nih.gov/clinvar/) variants represents a potential confounder in that we may be overestimating the frequency of disease relevant single *PARK2* mutations. Limiting our analyses to ClinVar variants only did not result in considerably different risk estimates across all comparisons (All studies, OR=1.70, P=2.65E-07; Biallelic studies, OR=1.99, P=0.036; non-EOPD studies, OR=1.55, P=5.6E-05). Another confounder relates to the fact that we observed a significant rate of occult second pathogenic mutations in putative monoallelic cases in our NeuroX cohort. The detected rate of occult biallelic carrier status was high in our two datasets (6/28, 21% and 4/9, 44%), approaching one half of PD cases with apparent monoallelic status. Additionally, several studies included in the analyses here have not searched for potentially hidden biallelic mutations in all cases and controls or have only interrogated a subset of *PARK2* mutations. Inclusion of these PD cases in our analysis is likely to appreciably influence our estimate. However, restricting the meta-analyses to 9 cohorts that searched for biallelic *PARK2* mutations in all cases and controls demonstrated that single mutations confer a 2-fold increase in risk in carriers. A further confounding factor is the use of EOPD cases (<50 years) in such studies which may be additionally inflating risk estimates as *PARK2* mutations are more likely to occur in PD cases of younger onset. This was observed in the IPDGC cohorts, with a higher estimate in the exome cohort compared to the NeuroX and resequencing cohorts. The additional removal of predominantly EOPD studies resulted in the loss of the original association (OR=1.23; P=0.614) but this was based on a few small studies (n=4). This suggests that current estimates of the effect of single mutations in modulating PD risk may not be accurate, but also stresses the importance of comprehensively searching for biallelic mutations in systematically recruited cohorts. It remains possible that there are further “occult” non-coding mutations affecting the promotor or splicing that have not yet been identified.

There are some limitations to our study. NeuroX biallelic cases will have been missed as not all possible PD-causing variants are represented on the chip (33). The same applies to the detection of biallelic carriers in the UK Biobank genotyping cohort. Identifying *PARK2* CNVs from NeuroX SNP genotype data using PennCNV may have missed smaller deletions/duplications. The false positive rate of PennCNV as a method for CNV detection was estimated to be 9.0-17.7%, with false positive CNVs predominantly small in size and occurring regardless of genotyping chip used (34). Our CNV detection false-positive rate in the NeuroX cohort is 6.9%. However, the fact that (i) the NeuroX variants are not evenly distributed across the *PARK2* locus (accounts for four misclassified samples), and (ii) we were looking for CNVs as small as a single exon, may have resulted in our approach missing or inaccurately calling CNVs in our large cohort comprising predominantly late-onset PD cases. There are limitations in defining CNVs from IPDGC and UK Biobank exome data, so CNVs were only investigated using MLPA in identified PD-Monoallelic exome cases. As the exome cohort predominantly consists of EOPD cases, it is likely that additional *PARK2* CNVs carriers were undetected. We therefore sought to validate the monoallelic status of available carriers by accessing diagnostic reports or directly assessing CNVs using MLPA and discovered a high rate of undetected second hits in both our datasets. Previous studies which have not systematically searched for second hits may have therefore erroneously determined the monoallelic carrier rate meaning that the estimates derived from our in-house cohorts and other published meta-analyses may not be accurate. It is therefore very important that any proposed increased risk associated with single *PARK2* mutations be considered with caution as, based on findings presented here, a substantial part of the reported excess on monoallelic carriers may relate to occult biallelic status.

In conclusion, while much of the data demonstrates that harbouring a single heterozygous *PARK2* mutation increases PD risk and that single *PARK2* CNVs may be more pathogenic than *PARK2* SNVs, there may be confounding factors. This is supported by our finding of no increased risk associated with single *PARK2* mutations upon restricting our analysis to studies that assessed biallelic mutations in cases and controls, and studies that did not include predominantly EOPD cases. Before the risk associated with single heterozygous mutations can be accurately defined, we highlight the importance of assessing ‘second hits’ in all cases and controls where both SNVs and CNVs are systematically interrogated in large-scale cohorts that have been systematically recruited.

## METHODS AND MATERIALS

High-quality genotype data from the NeuroX chip on 6,558 PD cases and 5,693 controls was assessed as part of the IPDGC (dbGaP Study Accession number: phs000918.v1.p1). Sample collection and variant genotyping have been described elsewhere (33). IPDGC exome sequencing data from 1,235 PD cases and 473 controls were used as a replication cohort (EGA Study Accession numbers: EGAS00001002103, EGAS00001002110, EGAS00001002113, EGAS00001002156; dbGaP Study Accession number: phs001103.v1.p1), and is described elsewhere (8). Additional replication cohorts used include: IPDGC resequencing cohort (cases=3,071, controls=2,162), UK Biobank Genotyping (cases=1,428, controls=312,098; downloaded April 2018, under application number 33601) and UK Biobank exome sequencing (cases=114, controls=38,263; downloaded May 2019) cohorts and the PPMI Exome sequencing *de novo* cohort (cases=385, controls=179). Duplicate samples were removed, where possible, from all analyses. Samples with missing call rates >5% were excluded during quality control. Variants (excluding synonymous) from known Mendelian PD-causing genes were extracted. Pathogenic mutations were identified as previously described (8). Rare *PARK2* (NM_013988 and NM_004562) CNVs were identified in the NeuroX cohort using PennCNV (34). CNVs spanning a minimum of ten variants were selected and visually confirmed. Monoallelic PD cases were defined as those carrying a single heterozygous pathogenic *PARK2* allele as defined according to OMIM (http://omim.org/), the Movement Disorder Society Genetic mutation database (https://www.mdsgene.org/) or the Parkinson Disease Mutation Database (http://www.molgen.vib-ua.be/PDMutDB/) (Supplementary Table 4). Where available, samples were investigated by (i) accessing sample diagnostic records, or (ii) using MLPA (SALSA P051 v.D1 probe mix [MRC-Holland, The Netherlands]) to confirm their monoallelic status. Without phasing information, any two PD-causing hits identified in an individual are assumed to be in trans.

To assess whether PD risk might be associated with (i) all monoallelic variants, (ii) CNVs alone or (iii) SNVs alone, as indicated by case-control differences, we used logistic regression correcting for gender and principal components (C1-4). Linear regression was used to investigate the impact of single AR mutations on AAO.

A literature review was undertaken (on 01/10/2019) to identify published data on heterozygous *PARK2* mutations, using search terms including combinations of the following terms: Parkinson’s disease, PD, *Parkin, PARK2* and heterozygous. Additional studies were identified by manual search of references cited in published articles. Should any of the studies include previously published data, the most recent data was selected where possible. Meta-analysis was conducted using standard methods modelling fixed effects, using Cochran’s Q-statistic to test for heterogeneity (P_het_) (35) and the I^2^ statistic (36) to quantify the proportion of the total variation caused by heterogeneity relating to possible differences in sample recruitment and assessment between studies. Meta-analyses were performed for CNVs and SNVs separately to investigate potential different risk profiles for each mutation type.

## Data Availability

For publicly available datasets, the data are available through application at the relevant sites.

## ACKNOWLEDGEMENTS

We would like to thank all of the subjects who donated their time and biological samples to be a part of this study. We also would like to thank all members of the International Parkinson Disease Genomics Consortium (IPDGC). See for a complete overview of members, acknowledgements and funding http://pdgenetics.org/partners.

This study was supported by Parkinson’s UK (grants 8047 and J-0804) and the Medical Research Council (G0700943 and G1100643). This work was supported in part by the Intramural Research Programs of the National Institute of Neurological Disorders and Stroke (NINDS), the National Institute on Aging (NIA), and the National Institute of Environmental Health Sciences both part of the National Institutes of Health, Department of Health and Human Services; project numbers 1ZIA-NS003154, Z01-AG000949-02 and Z01-ES101986. This research has been conducted using the UK Biobank Resource. In addition, this work was supported by the Department of Defense (award W81XWH-09-2-0128), and The Michael J Fox Foundation for Parkinson’s Research. This work was supported by National Institutes of Health grants R01NS037167, R01CA141668, P50NS071674, American Parkinson Disease Association (APDA); Barnes Jewish Hospital Foundation; Greater St Louis Chapter of the APDA. The KORA (Cooperative Research in the Region of Augsburg) research platform was started and financed by the Forschungszentrum für Umwelt und Gesundheit, which is funded by the German Federal Ministry of Education, Science, Research, and Technology and by the State of Bavaria. This study was also funded by the German Federal Ministry of Education and Research (BMBF) under the funding code 031A430A, the EU Joint Programme - Neurodegenerative Diseases Research (JPND) project under the aegis of JPND -www.jpnd.eu- through Germany, BMBF, funding code 01ED1406 and iMed - the Helmholtz Initiative on Personalized Medicine. This study is funded by the German National Foundation grant (DFG SH599/6-1) (grant to M.S), Michael J Fox Foundation, and MSA Coalition, USA (to M.S). The French GWAS work was supported by the French National Agency of Research (ANR-08-MNP-012). This study was also funded by France-Parkinson Association, Fondation de France, the French program “Investissements d’avenir” funding (ANR-10-IAIHU-06) and a grant from Assistance Publique-Hôpitaux de Paris (PHRC, AOR-08010) for the French clinical data. This study was also sponsored by the Landspitali University Hospital Research Fund (grant to SSv); Icelandic Research Council (grant to SSv); and European Community Framework Programme 7, People Programme, and IAPP on novel genetic and phenotypic markers of Parkinson’s disease and Essential Tremor (MarkMD), contract number PIAP-GA-2008-230596 MarkMD (to HP and JHu). Institutional research funding IUT20-46 was received of the Estonian Ministry of Education and Research (SK). The McGill study was funded by the Michael J. Fox Foundation and the Canadian Consortium on Neurodegeneration in Aging (CCNA). This study utilized the high-performance computational capabilities of the Biowulf Linux cluster at the National Institutes of Health, Bethesda, Md. (http://biowulf.nih.gov), and DNA panels, samples, and clinical data from the National Institute of Neurological Disorders and Stroke Human Genetics Resource Center DNA and Cell Line Repository. People who contributed samples are acknowledged in descriptions of every panel on the repository website. We thank the French Parkinson’s Disease Genetics Study Group and the Drug Interaction with genes (DIGPD) study group: Y Agid, M Anheim, F Artaud, A-M Bonnet, C Bonnet, F Bourdain, J-P Brandel, C Brefel-Courbon, M Borg, A Brice, E Broussolle, F Cormier-Dequaire, J-C Corvol, P Damier, B Debilly, B Degos, P Derkinderen, A Destée, A Dürr, F Durif, A Elbaz, D Grabli, A Hartmann, S Klebe, P. Krack, J Kraemmer, S Leder, S Lesage, R Levy, E Lohmann, L Lacomblez, G Mangone, L-L Mariani, A-R Marques, M Martinez, V Mesnage, J Muellner, F Ory-Magne, F Pico, V Planté-Bordeneuve, P Pollak, O Rascol, K Tahiri, F Tison, C Tranchant, E Roze, M Tir, M Vérin, F Viallet, M Vidailhet, A You. We also thank the members of the French 3C Consortium: A Alpérovitch, C Berr, C Tzourio, and P Amouyel for allowing us to use part of the 3C cohort, and D Zelenika for support in generating the genome-wide molecular data. We thank P Tienari (Molecular Neurology Programme, Biomedicum, University of Helsinki), T Peuralinna (Department of Neurology, Helsinki University Central Hospital), L Myllykangas (Folkhalsan Institute of Genetics and Department of Pathology, University of Helsinki), and R Sulkava (Department of Public Health and General Practice Division of Geriatrics, University of Eastern Finland) for the Finnish controls (Vantaa85+ GWAS data). We used genome-wide association data generated by the Wellcome Trust Case-Control Consortium 2 (WTCCC2) from UK patients with Parkinson’s disease and UK control individuals from the 1958 Birth Cohort and National Blood Service. Genotyping of UK replication cases on ImmunoChip was part of the WTCCC2 project, which was funded by the Wellcome Trust (083948/Z/07/Z). UK population control data was made available through WTCCC1. This study was supported by the Medical Research Council and Wellcome Trust disease centre (grant WT089698/Z/09/Z to NW, JHa, and ASc). As with previous IPDGC efforts, this study makes use of data generated by the Wellcome Trust Case-Control Consortium. A full list of the investigators who contributed to the generation of the data is available from www.wtccc.org.uk. Funding for the project was provided by the Wellcome Trust under award 076113, 085475 and 090355. Sequencing and genotyping done in McGill University was supported by grants from the Michael J. Fox Foundation, the Canadian Consortium on Neurodegeneration in Aging (CCNA), the Canada First Research Excellence Fund (CFREF), awarded to McGill University for the Healthy Brains for Healthy Lives (HBHL) program and Parkinson’s Society Canada. We thank Jeffrey Barrett and Jason Downing (Illumina Inc) for assistance with the design of the ImmunoChip and NeuroX arrays. DNA extraction work that was done in the UK was undertaken at University College London Hospitals, University College London, who received a proportion of funding from the Department of Health’s National Institute for Health Research Biomedical Research Centres funding. This study was supported in part by the Wellcome Trust/Medical Research Council Joint Call in Neurodegeneration award (WT089698) to the Parkinson’s Disease Consortium (UKPDC), whose members are from the UCL Institute of Neurology, University of Sheffield, and the Medical Research Council Protein Phosphorylation Unit at the University of Dundee. We thank the Quebec Parkinson’s Network (http://rpq-qpn.org) and its members. This work was supported by the Medical Research Council grant MR/N026004/1. The Braineac project was supported by the MRC through the MRC Sudden Death Brain Bank Grant (MR/G0901254) to J.H. P.A.L. was supported by the MRC (grants MR/N026004/1 and MR/L010933/1) and Michael J. Fox Foundation for Parkinson’s Research. This work was supported by the Simpson Querrey Center for Neurogenetics (to DK).

## ABBREVIATIONS

%: Percent
AAO: Age at onset
AR: Autosomal recessive
c.: coding DNA reference sequence
C1-4: Principal component 1 to 4
CI: Confidence interval
CNV: Copy number variant
Coeff: β-Coefficient
dbGaP: Database of Genotypes and Phenotypes
DNA: Deoxyribonucleic acid
EGA: European Genome-phenome Archive
EOPD: Early-onset Parkinson’s
*FBXO7*: F-box protein 7
Freq: Frequency
I^2^: Proportion of total variation caused by heterogeneity
IPDGC: International Parkinson’s Disease Genomics Consortium
*LRRK2*: Leucine-rich repeat kinase 2
MLPA: Multiplex ligation-dependent probe amplification
MRC-Holland: Microbiology Research Centre Holland
N: Number
NM_*: Messenger RNA sequence identifier
OMIM: Online Mendelian Inheritance in Man
OR: Odds ratio
P: P-value
p.: protein reference sequence
*PARK2*: Parkin
*PARK7*: Parkinsonism associated deglycase or *DJ-1*
PD: Parkinson’s
PD-Monoallelic: Parkinson’s cases harbouring a single heterozygous *PARK2* mutation
P_het_: Cochran’s Q-statistic test for heterogeneity P-value
*PINK1*: PTEN-Induced putative kinase 1
P_log_: Logistic regression P-value
PPMI: Parkinson’s Progression Markers Initiative
P_reg_: Linear regression P-value
Reseq: Resequencing
*SNCA*: α-Synuclein
SNV: Single nucleotide variant
*VPS35*: Vacuolar protein sorting 35, yeast, homolog of
*vs*.: Versus

